# Longitudinal gray matter trajectories and cognitive performance during rehabilitation after moderate–severe traumatic brain injury: a longitudinal VBM pilot study

**DOI:** 10.64898/2026.07.06.26357170

**Authors:** Rhideeta Jalal, Jonathan Yoon, Jessica Ashley, Mark Ashley, Grace Griesbach, Brenda Bartnik-Olson

## Abstract

Moderate-to-severe traumatic brain injury (msTBI) is recognized as a chronic and evolving neurological condition characterized by progressive structural brain changes and persistent cognitive impairment. While prior studies have demonstrated widespread atrophy following msTBI, less is known regarding the longitudinal trajectory of gray matter (GM) changes during recovery and post-rehabilitation. The current study used longitudinal voxel-based morphometry (VBM) to characterize GM volume changes over a period of 7 - 12 months, in individuals with msTBI relative to healthy controls (HC). Associations between regional GM volume and neuropsychological functioning were examined. Twenty-eight participants (14 msTBI, 14 HC) completed MRI and neuropsychological assessments across three timepoints spanning outpatient rehabilitation and follow-up. Longitudinal VBM analyses revealed significant group and time interactions within subcortical and limbic regions. Relative to HC, individuals with msTBI showed lower GM volume in these regions at baseline, with trajectories that converged toward HC values (right hippocampus) or increased relative to HC over the rehabilitation period (bilateral pulvinar), whereas the right amygdala and inferior cerebellar vermis remained persistently reduced. Significant longitudinal improvements in memory and psychomotor speed during the rehabilitation period were demonstrated in msTBI. Greater (preserved) GM volume within the right hippocampus, thalamus, and bilateral pulvinar was associated with better performance across measures of verbal memory, processing speed, executive functioning, and cognitive flexibility. These findings suggest that msTBI is associated with dynamic structural brain changes involving subcortical, limbic, and cerebellar networks, and that the rehabilitation period was accompanied by relative volumetric stabilization in these regions and by meaningful cognitive improvement.

## 1. Introduction

Traumatic brain injury (TBI) is a leading cause of long-term neurological disability worldwide, affecting an estimated 69 million individuals annually (Dewan et al., 2019; GBD 2016 Traumatic Brain Injury and Spinal Cord Injury Collaborators, 2019; Khattak et al., 2026). TBI is associated with substantial adverse health, social, and economic outcomes (Faul & Coronado, 2015; Humphreys et al., 2013; Wilson et al., 2017) and has been linked to an increased risk of later-life neurodegenerative disease (Shively et al., 2012; Wilson et al., 2017; Graham & Sharp, 2019). TBI exists along a continuum of severity, ranging from mild, to moderate, and severe, with individuals frequently experiencing impairments in attention, processing speed, executive functioning, emotional regulation, and memory. Mild TBI (mTBI) is typically associated with transient symptoms and recovery within weeks to months (McCrory et al., 2001), however, approximately 15-30% of individuals experience persistent symptoms, particularly a decline of neurocognitive function, physical function, and develop increased mood disorders that persist for years post injury (McInnes et al., 2017; Shenton et al., 2012). In contrast, moderate-to-severe TBI (msTBI), defined by loss of consciousness greater than 30 minutes and/or post-traumatic amnesia greater than 24 hours, often results in permanent cognitive, emotional, and functional impairments that can significantly impact quality of life and independence (Maas et al., 2008; Wilson et al., 2017); (Marino et al., 2025).

Structural neuroimaging has played a central role in characterizing the neural consequences of TBI. Cross-sectional studies using voxel-based morphometry (VBM), cortical thickness analysis, and volumetric MRI measurements have reported widespread gray matter volume loss, particularly in the frontal and temporal cortex, hippocampi, and subcortical structures including the thalami (Cole et al., 2018; Ng et al., 2008). While traditionally TBI has been conceptualized as an acute neurological insult, there is increasing evidence that TBI represents a chronic and evolving condition, characterized by progressive structural brain changes and long-term neurodegenerative processes (Smith et al., 2013); (Graham & Sharp, 2019). Despite the generally consistent findings, cross-sectional studies are limited in their ability to capture dynamic changes and their relationship to the cognitive and functional consequences of msTBI. In contrast, longitudinal study designs have become increasingly used to address these limitations and improve sensitivity by reducing inter-subject variability (Ashburner & Ridgway, 2012).

A substantial body of longitudinal neuroimaging research suggests that msTBI has been associated with accelerated brain atrophy relative to normal aging, with a loss of brain tissue that can occur at a rate approximated around 5% per year compared to normal age-related decline of 0.2-0.5% (Harris et al., 2019). Similarly, Cole et al., (2018) demonstrated that moderate-severe TBI patients have an accelerated neurodegenerative rate with a mean predicted age difference between chronological and estimated brain age of 4.66 years for gray matter (GM) and 5.97 years for white matter (WM; Cole et al., 2015). The study also reported a strong correlation between chronicity of injury and brain atrophy, suggesting brain tissue loss increases during the chronic post-injury period. Similarly, other investigations report acute brain tissue losses ranging from 3-8.4% within the first 6-8 months post-injury, with progressive atrophy continuing into the chronic phase (Ding et al., 2008; Warner et al., 2010).

General cortical atrophy is a common consequence of msTBI (Ding et al., 2008; Harris et al., 2019; Warner et al., 2010) with the bulk loss occurring in the first year (Sidaros et al., 2009). Whole brain volume loss, notably in frontal, temporal, and occipital cortices was found to significantly lower at 1-year post-injury compared to controls (Cole et al., 2018). Several studies have also examined regional volume loss, finding significant volume reduction in subcortical regions including the thalamus, putamen, caudate, and amygdala compared to controls (Cole et al., 2018; Zagorchev et al., 2016). The thalamus has emerged as one of the most consistently implicated subcortical structures in TBI (Cole et al., 2018; Lutkenhoff et al., 2013; Marcuse et al., 2025), given its role as a central relay station and integrative hub within cortico-subcortical networks, thalamic injury may have widespread effects on attention, processing speed, and executive functioning. The amygdala and cerebellum have also been implicated in TBI, given their roles in emotion regulation, affective control, coordination, motor control (Bigler, 2023; McCorkle et al., 2021). Hippocampal volume loss has been shown to occur between 2 scan points in TBI compared to controls (Kriukova et al., 2024; Ng et al., 2008), and is increasingly recognized as relevant to post-TBI outcomes, given its vulnerability to hypoxia, as well as its central role in memory encoding and retrieval.

Few longitudinal studies have characterized the regional pattern of gray matter (GM) change over time in msTBI relative to controls or examined how these changes relate to cognitive functioning and quality of life. A detailed understanding of the relationship between TBI and its neural consequences, including cortical and subcortical changes, cognitive impairment, and functional outcomes is critical for alleviating the emotional, social, and economic burden of TBI. The purpose of this pilot study was to characterize the pattern and scale of GM volume change occurring during a period of 7 – 12 months, which spanned a period of outpatient rehabilitation. Using a longitudinal VBM approach, we characterize the pattern of GM change and examine the relationship between the GM volume changes and measures of cognitive performance (e.g., memory, attention) and quality of life to establish preliminary insight into the neuroanatomical response to standard outpatient rehabilitation. We predicted significant GM alterations in the msTBI participants, compared to healthy controls (HC), particularly within subcortical and medial temporal regions, despite the small sample size. Because the imaging window spanned active rehabilitation, whether these regions would show continued decline or relative stabilization over time was treated as an exploratory question. Additionally, we hypothesized that greater GM volume would be associated with better cognitive performance and quality of life outcomes in both groups.

## 2. Methods

### 2.1 Study participants and data acquisition

Imaging and neuropsychological data used in this study were acquired as part of a longitudinal multimodal imaging study investigating the structural correlates of recovery following a msTBI and participation in a full-time rehabilitation program either as a resident or day treatment. This program involved participation of up to 5 days a week, 6 hours per day. Participants with msTBI were enrolled based on the following inclusion criteria: had sustained a TBI, were currently enrolled in an inpatient or day treatment program at the Center for Neuro Skills for a minimum of 4 weeks, between the ages of 18 - 60 years, had at least an 8th grade education, could read and speak English, and had the ability to comply with the research protocol (behaviorally, psychologically, and intellectually). Twenty individuals with msTBI provided informed consent and underwent baseline neuropsychological evaluation and MR imaging. On average, the first imaging time point was performed at 9.0 ± 8.6 months post-injury.

Follow-up MR imaging and neuropsychological testing was performed on average, 3 months (scan 2) and on average, 9 months (scan 3), following the enrollment (Table 1). Sixty-four percent of the msTBI participants had a 3rd scan at a mean of 20.6 ± 10.3 months post-injury. HC participants were recruited from the surrounding community and used to identify GM volume changes due to injury. The exclusion criteria for controls were 1) current substance abuse as determined by family/ informant interview, 2) having a contraindication to MRI such as pregnancy, surgical clips, metallic artificial prostheses, surgically implanted pacemakers, ventriculoperitoneal shunt or claustrophobia, 3) medical history of neurological disease including psychiatric history of schizophrenia, intellectual disability, and/or a diagnosis of Alzheimer’s disease, 4) prior TBI (hospitalization required), and 5) TBI resulting from penetrating head wound or blast injury. HC participants were scanned at comparable time intervals. All imaging data underwent quality control procedures, including visual inspection for artifacts, segmentation quality, and motion artifacts. Following exclusion of participants with incomplete data (less than 2 MRI exams or neuropsychological evaluations) or poor image quality, a total of 28 participants (msTBI= 14, HC = 14) were included in the final analysis. All participants provided written informed consent, and the study was approved by Advarra Institutional Review Board (approval number 00017659) and is compliant with all relevant codes and legislation. Demographic and clinical information for the final sample can be found in Table 1.

**Table 1.**
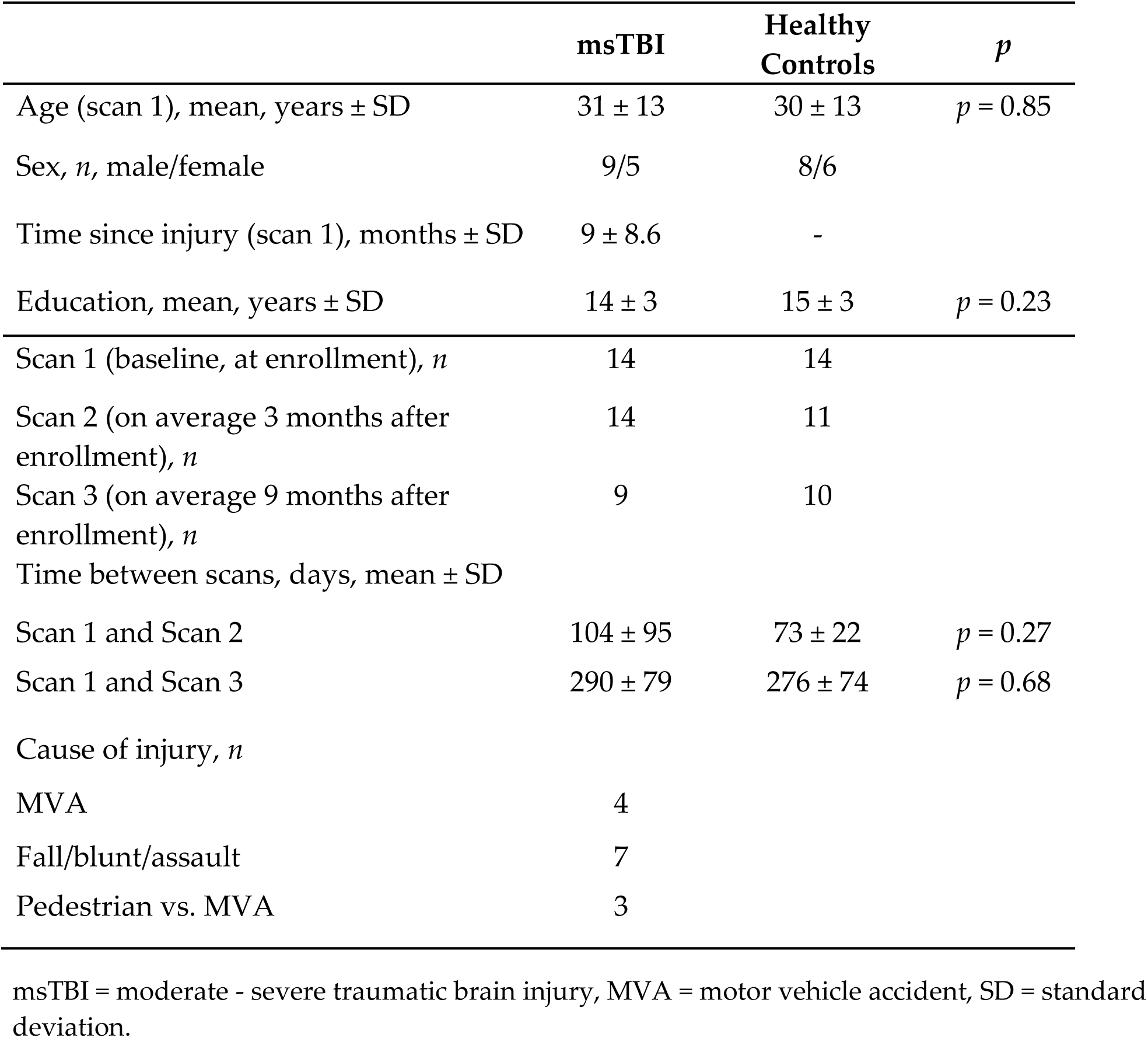
Demographic and injury characteristics of the msTBI and health control participants.

|  | **msTBI** | **Healthy Controls** | ***p*** |
| --- | --- | --- | --- |
| Age (scan 1), mean, years ± SD | 31 ± 13 | 30 ± 13 | $p = 0.85$ |
| Sex, *n*, male/female | 9/5 | 8/6 |  |
| Time since injury (scan 1), months ± SD | 9 ± 8.6 | - | |
| Education, mean, years ± SD | 14 ± 3 | 15 ± 3 | $p = 0.23$ |
| Scan 1 (baseline, at enrollment), *n* | 14 | 14 |  |
| Scan 2 (on average 3 months after enrollment), *n* | 14 | 11 |  |
| Scan 3 (on average 9 months after enrollment), *n* | 9 | 10 |  |
| Time between scans, days, mean ± SD | | | |
| Scan 1 and Scan 2 | 104 ± 95 | 73 ± 22 | $p = 0.27$ |
| Scan 1 and Scan 3 | 290 ± 79 | 276 ± 74 | $p = 0.68$ |
| Cause of injury, *n* |  |  |  |
| MVA | 4 |  |  |
| Fall/blunt/assault | 7 |  |  |
| Pedestrian vs. MVA | 3 |  |  |

msTBI = moderate - severe traumatic brain injury, MVA = motor vehicle accident, SD = standard deviation.

### 2.2 Neuroimaging acquisition

MRI data was acquired on a Siemens 3-Tesla (3T) scanner using a 32-channel head coil. High resolution structural images were acquired using a T1-weighted magnetization-prepared rapid gradient echo (MPRAGE) sequence with TR = 2400 ms, TE = 2.2 ms, FOV = 320 x 208 mm, and 0.8 mm isotropic voxel size. T1-weighted images were used in the analysis due to the high gray-white matter contrast.

### 2.3 Neuroimaging data processing

Voxel based morphometry (VBM) was performed using the Computational Anatomy Toolbox (CAT12 toolbox, v. 19) for Statistical Parametric Mapping (SPM, v. 12) deployed in a Matlab environment (v. 2023, Mathworks Inc. USA). Longitudinal preprocessing was performed using the CAT12 longitudinal segmentation pipeline, using the neuroplasticity option. Images were segmented into GM, WM, and cerebrospinal fluid (CSF). The diffeomorphic anatomic registration via exponential like algebra algorithm (DARTEL) tool was used to normalize the segmented GM and WM volumes to the MNI152 standard template. After affine and non-linear registration, images were modulated using Jacobian determinants and smoothed with an 8mm full width half maximum Gaussian kernel.

### 2.4 Neuroimaging Analysis

Neuroimaging analysis consisted of cross-sectional comparisons of global and regional brain volumes between msTBI participants and controls at each of the 3 individual scan time points, longitudinal analyses of changes in tissue volumes over the 7 – 12 month study period, and a region of interest (ROI)-based analysis.

#### 2.4.1 Longitudinal VBM analyses

Voxel-wise longitudinal analyses were performed in SPM12 using a flexible factorial model implemented on the longitudinally realigned and modulated GM maps generated by the CAT12 longitudinal processing pipeline. The model included 3 factors: subject, group (msTBI vs HC), and time (scan 1, scan 2, and scan 3). Subject was specified as a repeated-measures factor (independence = no, variance = unequal) to account for within-subject correlations across the longitudinal observations. Group was specified as the between-subjects factor (independence = yes, variance = unequal) and time was specified as a within-subject factor (independence = no, variance = equal). Total intracranial volume (TIV) was used as a covariate (centering = overall mean) to control for differences in head size. Additional covariates included age (centering = overall mean) and sex. Unequal numbers of subjects across time were accommodated within the flexible factorial model without requiring the exclusion of participants with incomplete observations. An implicit mask with an absolute threshold of 0.1 was used to ensure that only GM voxels with intensity value of 0.1 or higher were included in the analyses. The flexible factorial model computed three statistical contrasts: (a) main effect of time, (b) group difference between msTBI and HC GM volume at scan 1, scan 2, and scan 3, and (c) the interaction of group and time. Voxel wise statistical significance was assessed using a family-wise error (FWE)-corrected threshold of *p* < .05, with an initial voxel-wise threshold of *p* < .001 and a cluster forming threshold of k ≥ 50 voxels.

#### 2.4.2 Cross-sectional VBM analyses

Regions of suprathreshold clusters of significant voxel-wise differences in GM were identified using the Neuromorphometrics atlas enabled in CAT12. Group-by-time effects on GM volumetric measurements within these regions were assessed using repeated-measures ANOVA, with group (msTBI vs HC) as the between-subjects factor and timepoint as the within-subjects factor. Significant main and interaction effects were conducted with post-hoc independent-samples t-tests at individual scan timepoints. Statistical significance was set at *p* < .05 after Bonferroni-Holm correction for multiple comparisons.

### 2.5 Neuropsychological testing

Cognitive functioning was assessed at each imaging timepoint using a standardized battery selected to assess cognitive domains commonly affected by TBI, including measures of attention, processing speed, executive function, learning and memory. Patient-reported measures of emotional and functional outcome were also assessed.

Cognitive performance was evaluated using the CNS Vital Signs (CNSVS) computerized neurocognitive battery, which provides norm-referenced standard scores across multiple domains. Domains assessed included verbal memory, visual memory, psychomotor speed, processing speed, complex attention, cognitive flexibility, and executive function. Performance across domains were measured using age-standardized composite scores generated by the CNS Vital Signs battery. Verbal episodic memory was assessed using the CVLT with performance measured using standardized scaled scores, with higher scores indicating better verbal memory performance. Mood symptoms were assessed using the BDI-II, a self-report measure assessing cognitive, affective, and somatic symptoms of depression. Total raw scores range from 0-63, with higher scores indicating greater depression symptom severity. Additional measures included the Neuro-QoL (quality of life in neurological disorders) domains of anxiety, fatigue, and satisfaction with social roles and activities, and sleep disturbance. Neuro-QoL domains were measured as T-scores, for which higher scores on anxiety, fatigue, and sleep disturbance are worse. Higher scores on satisfaction with social roles and activities is better.

### 2.6 Within-group differences in neuropsychological measures in msTBI

To examine the longitudinal changes in neuropsychological functioning across the three assessment timepoints (T1, T2, and T3) within the msTBI group, linear mixed model (LMM) analyses were conducted for each neuropsychological measure using IBM SPSS Statistics (Version 31.0; IBM Corp., Armonk, NY, USA). Bonferroni-adjusted pairwise comparisons were performed to evaluate differences between timepoints. Statistical significance was set to a threshold of *p* < .05.

### 2.7 Association between GM volume, cognitive function, memory, and mood

To examine the functional relevance of the volumetric brain changes, correlation analyses were performed comparing the mean ROI-based GM volume and neuropsychological performance. To account for effects of age and global brain size, Pearson’s partial correlations were computed controlling for age and TIV.

## 3. Results

### 3.1 Global group differences in tissue volume

The percentage of global GM, WM, and CSF relative to the estimated TIV were compared between msTBI and HC participants at each imaging time point to categorize absolute group differences (Table 2). There were no differences in GM, WM, and CSF volume at any scan time point between the msTBI and HC groups, with tissue volumes in the msTBI group stable across the evaluation period.

**Table 2.** Global GM, WM, and CSF percentages for msTBI participants and healthy controls across scan 1, scan 2, and scan 3.

|  | msTBI |  |  | Healthy Controls |  |  |  |
| --- | --- | --- | --- | --- | --- | --- | --- |
|  | Scan 1 | Scan 2 | Scan 3 | Scan 1 | Scan 2 | Scan 3 | *p*-value |
| **GM** | 43.80 | 43.80 | 44.51 | 45.15 | 45.96 | 45.04 | n.s. |
| **(% of TIV)** | (3.90) | (3.23) | (1.90) | (2.95) | (2.76) | (1.91) |  |
| **WM** | 36.36 | 35.84 | 35.55 | 36.35 | 35.85 | 36.29 | n.s. |
| **(% of TIV)** | (2.23) | (2.13) | (1.83) | (1.66) | (1.52) | (1.59) |  |
| **CSF** | 18.49 | 16.66 | 18.66 | 19.84 | 20.37 | 19.92 | n.s. |
| **(% of TIV)** | (2.56) | (3.03) | (2.57) | (2.04) | (5.66) | (1.75) |  |

Values expressed as a percentage of the mean volume of the estimated total intracranial volume (standard deviation) to account for inter-individual differences in head size. CSF = cerebrospinal fluid; GM = gray matter; TIV = total intracranial volume; WM = white matter. P-values reflect between-group comparisons across the timepoints using ANCOVA, controlling for sex where $*p < 0.05$ .

### 3.2 Longitudinal volumetric analysis

A significant main effect of time was observed across widespread gray matter regions, with the strongest effects in the medial temporal GM regions including the right hippocampus and right amygdala averaging across both msTBI and HC groups (Figure 1; FWE correction *p* < .05; k > 50 voxels). Additionally, widespread significant group x time interactions were observed (FWE correction *p* < .05; k > 50 voxels) indicating widespread differences in the longitudinal GM trajectories between msTBI and HC subjects. Peak effects were noted in the subcortical, limbic, and medial temporal GM regions, including the pulvinar/posterior thalamus nuclei, right hippocampus, right amygdala, and cerebellar vermis regions. In the bilateral pulvinar/posterior thalamic nuclei, the GM trajectories diverge between the msTBI and HC at the first two scans, with msTBI participants showing an increase in GM over time, while HC pulvinar volumes remain stable (Figure 2). Similarly, GM volumes in the right hippocampus of msTBI participants are significantly lower than HC at scan 1 and 2 before converging with HC values at scan 3 (Figure 3). In contrast, the GM volume of the right amygdala and the inferior cerebellar vermis of msTBI participants was significantly lower than HC participants and remained so throughout the 7 - 12 month scan period (Figure 4 and 5). Across all three scan timepoints, there were no regions in which the msTBI group exhibited greater GM volume compared to HC. See Table 3 for mean regional GM volumes of the significant clusters.

**Figure 1.**
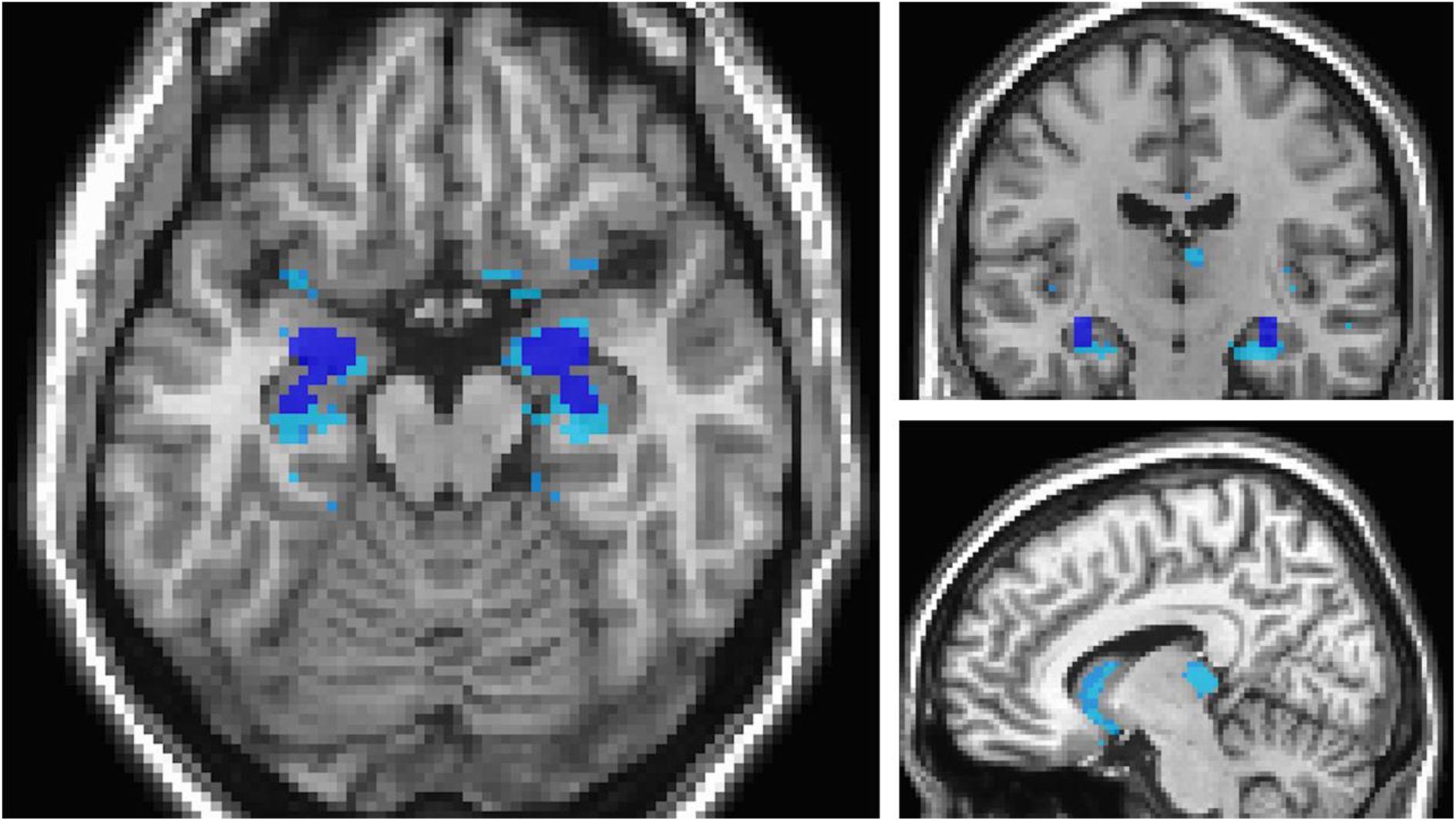
Effect of time on gray matter volume. Statistical parametric maps illustrate regions of significant group and time interactions between msTBI and healthy controls displayed on a MNI152 2mm standard image. Cool colors indicate regions where GM change differed significantly. Significant clusters were observed in the subcortical and limbic regions, including the thalamus, pulvinar nuclei, right hippocampus, right amygdala, and cerebellar regions. Statistical maps were set to a threshold of *p* < .001 with cluster-level family wise error (FWE) correction at *p* < .05 (k > 50 voxels). GM = gray matter, GMV = gray matter volume, msTBI = moderate-severe TBI.

**Figure 2.**
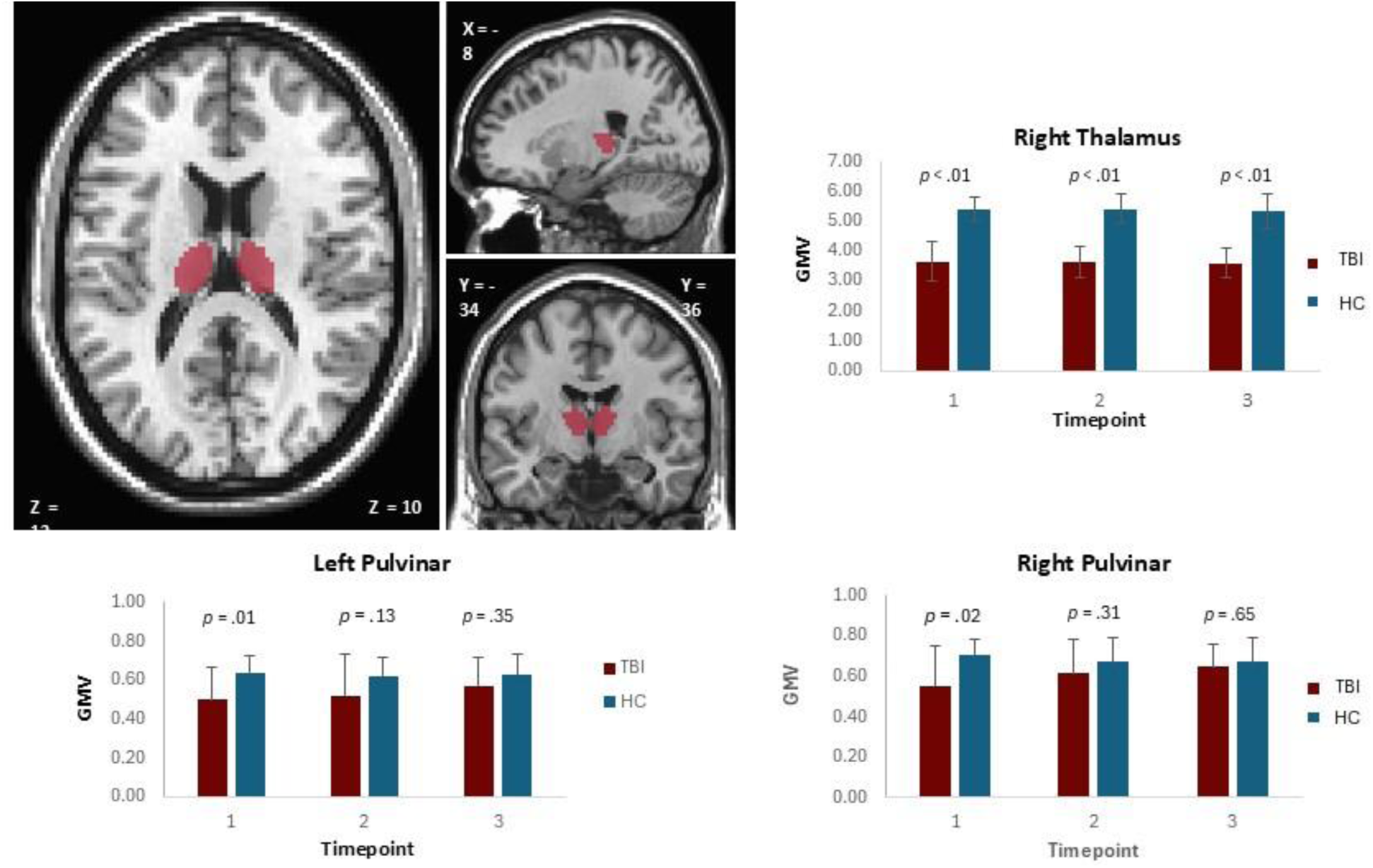
Effect of group and time on gray matter volume (cm^3^) in the bilateral pulvinar nuclei of the thalamus. GM trajectories diverge between the msTBI and HC at the first two scans, with msTBI participants showing an increase in GM over time, while HC pulvinar volumes remain stable. GMV in the right thalamus was significantly lower that HC participants at all 3 time points. Error bars represent standard deviation of the mean. Group differences were assessed using independent samples t-tests. Statistical significance was set at *p* < .05. GM = gray matter, GMV = gray matter volume, HC = healthy control, msTBI = moderate-severe TBI.

**Figure 3.**
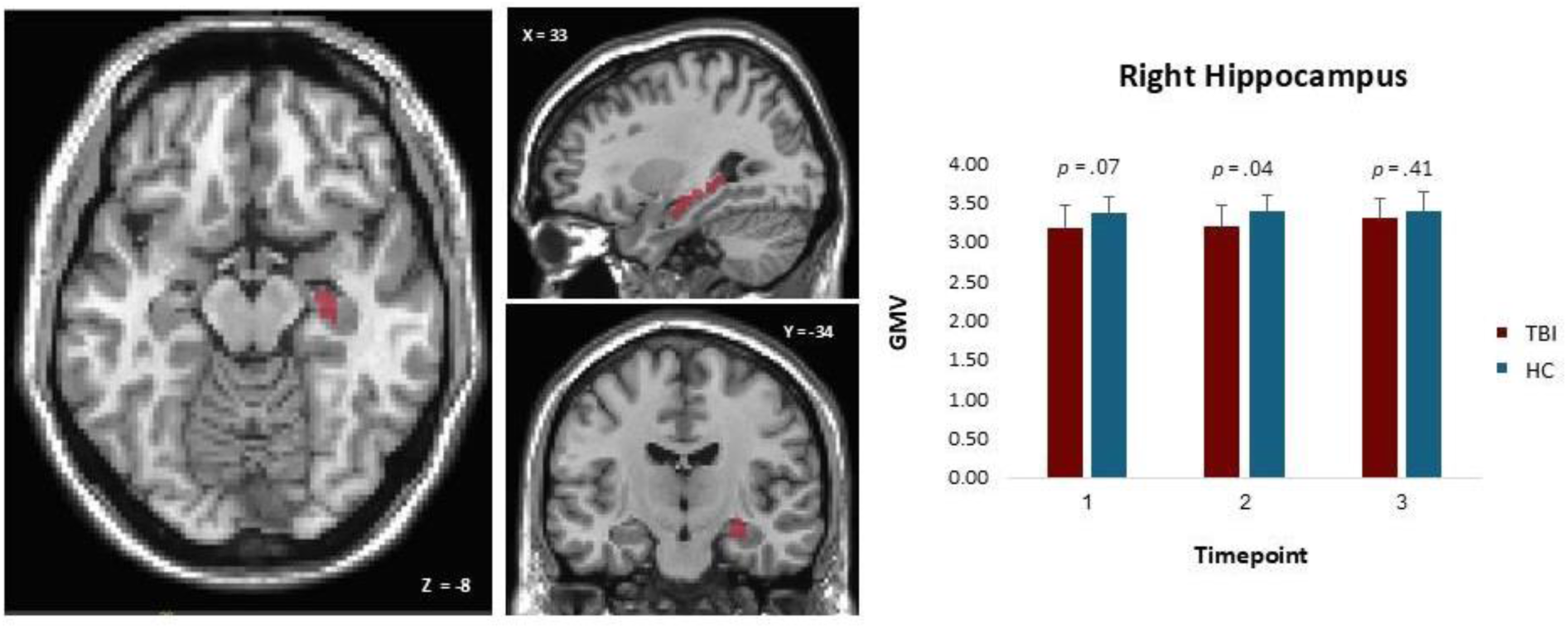
Effect of group and time on gray matter volume (cm^3^) in the right hippocampus. GM trajectories in the right hippocampus demonstrated group differences across longitudinal timepoints, with HC showing slightly greater GMV relative to the msTBI group at timepoint 2, *p* = .04. Error bars represent standard deviation of the mean. Group differences were assessed using independent samples t-tests. Statistical significance was set at *p* < .05. GM = gray matter, GMV = gray matter volume, HC = healthy control, msTBI = moderate-severe TBI.

**Figure 4.**
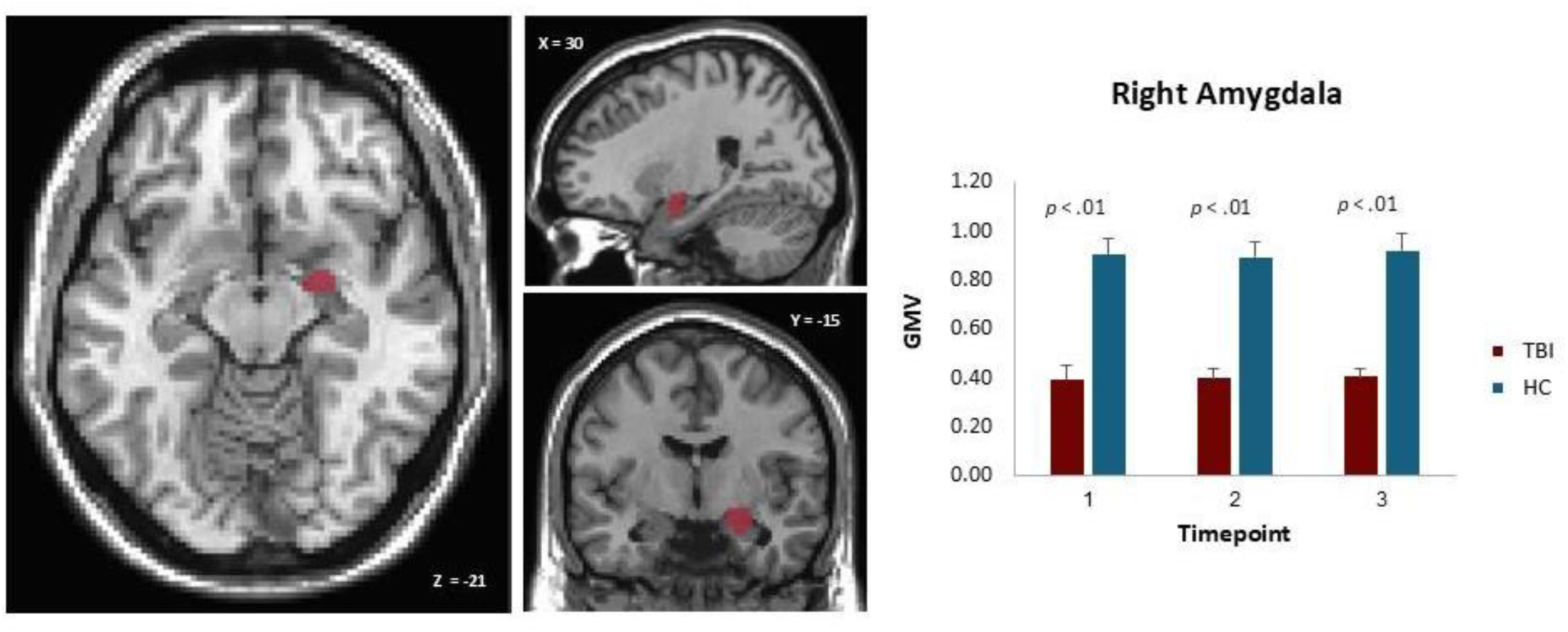
Effect of group and time on gray matter volume (cm^3^) in the right amygdala GM trajectories in the right amygdala demonstrated reduced GMV in the msTBI group relative to HC across all timepoints. Significant between-group differences were observed at timepoints 1, 2, and 3, *p* < .01. Error bars represent standard deviation of the mean. Group differences were assessed using independent samples t-tests. Statistical significance was set at *p* < .05. GM = gray matter, GMV = gray matter volume, HC = healthy control, msTBI = moderate-severe TBI.

**Figure 5.**
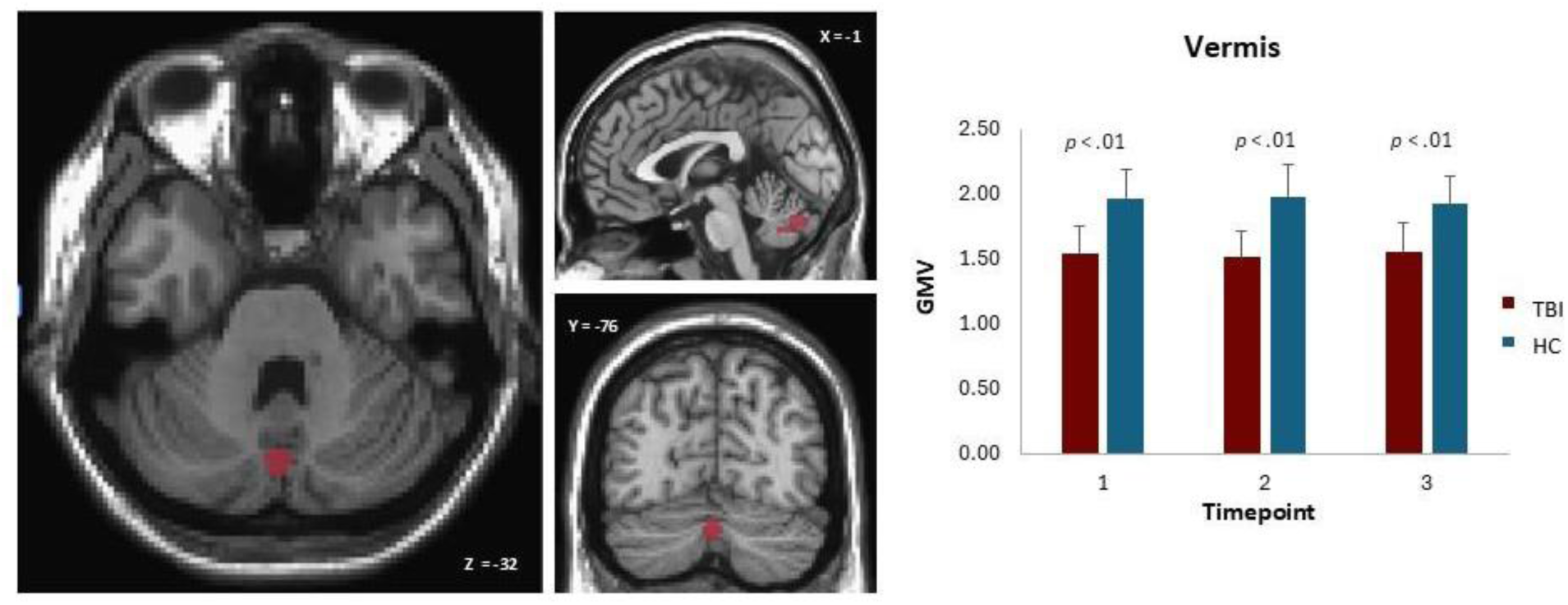
Effect of group and time on gray matter volume in the inferior cerebellar vermis. GM trajectories in the inferior cerebellar vermis demonstrated reduced GMV in the msTBI relative to HC across all timepoints. Significant between-group differences were observed at timepoints 1, 2, and 3, *p* < .01. Error bars represent standard deviation of the mean. Group differences were assessed using independent samples t-tests. Statistical significance was set at *p* < .05. GM = gray matter, GMV = gray matter volume, HC = healthy control, msTBI = moderate-severe TBI.

**Table 3.** Gray matter volume (cm^3^) of regions corresponding to significant clusters of longitudinal change in msTBI subjects compared to HC.

| Region | msTBI |  |  | Healthy Controls |  |  |
| --- | --- | --- | --- | --- | --- | --- |
|  | Scan 1 | Scan 2 | Scan 3 | Scan 1 | Scan 2 | Scan 3 |
| Right Hippocampus | 3.16<br>(0.30) | 3.18<br>(0.27) | 3.29<br>(0.25) | 3.36<br>(0.20) | 3.40<br>(0.19) | 3.39<br>(0.24) |
| Right Amygdala | 0.39<br>(0.05) | 0.40<br>(0.04) | 0.41<br>(0.03) | 0.90<br>(0.07) | 0.89<br>(0.06) | 0.92<br>(0.07) |
| Right Thalamus | 3.65<br>(0.67) | 3.63<br>(0.52) | 3.59<br>(0.49) | 5.38<br>(0.43) | 5.40<br>(0.50) | 5.34<br>(0.57) |
| Vermis | 1.55<br>(0.20) | 1.52<br>(0.21) | 1.57<br>(0.22) | 1.97<br>(0.22) | 1.99<br>(0.25) | 1.93<br>(0.21) |
| Left Pulvinar | 0.49<br>(0.16) | 0.51<br>(0.21) | 0.57<br>(0.14) | 0.63<br>(0.09) | 0.62<br>(0.10) | 0.63<br>(0.10) |
| Right Pulvinar | 0.55<br>(0.19) | 0.61<br>(0.17) | 0.65<br>(0.11) | 0.71<br>(0.07) | 0.67<br>(0.12) | 0.67<br>(0.12) |

Mean gray matter (GM) volumes (standard deviation) of regions corresponding to significant clusters of longitudinal change at scan 1 (baseline), scan 2 (on average 3-months), and scan 3 (on average 9-months) in moderate-to-severe traumatic brain injury (msTBI) compared to healthy controls (HC). Regions were defined using the Neuromorphometrics atlas.

### 3.3 Within-group differences of neuropsychological measures in msTBI

Significant main effects of time were observed for verbal memory, psychomotor speed, CVLT short-delay free recall, CVLT short-delay cued recall, CVLT long-delay free recall, CVLT long-delay cued recall, and satisfaction with social roles and activities in the msTBI participants. Specifically, verbal memory showed significant improvement across time F(2, 21.58) = 3.45, *p* = .05, with Bonferroni-corrected pairwise comparisons indicating higher performance at timepoint 3 relative to timepoint 1 (*p* = .05). Psychomotor speed also significantly improved across timepoints F(2, 19.43) = 5.08, *p* = .02, reflecting faster and improved performance at later timepoints relative to baseline, more specifically, between timepoint 2 relative to timepoint 1 (*p* = .03) and between timepoint 3 relative to timepoint 1 (*p* = .02). Similarly, significant longitudinal improvements were observed for CVLT performance, including short-delay free recall F(2, 19.59) = 4.96, *p* = .02, with Bonferroni-corrected pairwise comparisons indicating higher performance at timepoint 2 compared to timepoint 1 (*p* = .02). Short-delay cued recall showed significant improvement across time F(2, 19.67) = 5.76, *p* = .01, with Bonferroni-corrected pairwise comparisons indicating higher performance at timepoint 2 compared to timepoint 1 (*p* = .01). Long-delay free recall showed significant improvement across time F(2, 20.24) = 5.59, *p* = .01, with Bonferroni-corrected pairwise comparisons indicating higher performance at timepoint 2 compared to timepoint 1 (*p* = .01). Long-delay cued recall showed significant improvement across time F(2, 20.12) = 5.43, *p* = .01, reflecting improved performance at later timepoints relative to baseline, more specifically, between timepoint 2 relative to timepoint 1 (*p* = .02) and between timepoint 3 relative to timepoint 1 (*p* = .02). Lastly, satisfaction with social roles and activities also significantly changed across timepoints F(2, 20.08) = 4.70, *p* = .02, with Bonferroni-corrected pairwise comparisons indicating higher performance at timepoint 2 compared to timepoint 1 (*p* = .02).

### 3.4 Relationship between longitudinal gray matter volume changes and neuropsychological status in msTBI

Significant associations were observed between the longitudinal changes in GM volume and neuropsychological measures in msTBI participants.

#### 3.4.1 Baseline

At the initial evaluation, a positive correlation was observed between the right hippocampus GM volume and psychomotor speed (p = .02, r = .646), cognitive flexibility (p = .04, r = .591), processing speed (p = .03, r = .624), executive functioning (p = .04, r= .590), CVLT long-delay free recall (p = .05, r = .585), and CVLT long-delay cued recall (p = .03, r = .629), such that higher GM volumes were associated with better neuropsychological performance. A positive correlation was observed in the right amygdala GM volume with verbal memory performance, specifically, CVLT long-delay free recall (p = .05, r =.584), and long-delay cued recall (p = .03, r= .617). Similarly, a positive correlation was observed in the right thalamus GM volume with verbal memory performance, specifically, CVLT short-delay free recall (p = 02, r = .652), short-delay cued recall (p = 02, r= .662), long-delay free recall (p = 04, r =.590), and long-delay cued recall (p = .01, r =.699). A positive correlation was observed in the right pulvinar GM volume with higher CVLT short-delay free recall (p = 02, r =.643), short-delay cued recall (p = .03, r = .623), and long-delay cued recall (p = .01, r = .685). A positive correlation was observed in the left pulvinar GM volume with higher CVLT short-delay free recall (p = .05, r = .576), short-delay cued recall (p = .02, r =.666), long-delay free recall (p = .04, r = .599), and long-delay cued recall (p = .02, r =.660).

#### 3.4.2 Time 3

At the final evaluation, the positive correlation between the right thalamus GM volume and verbal memory performance remained, specifically, CVLT short-delay free recall (p = .03, r = .733), short-delay cued recall (p = .01, r = .795), long-delay free recall (p = .03, r = .722), and long-delay cued recall (p = .02, r = .739) were all associated with high thalamic GM volume.

## 4. Discussion

msTBI is increasingly recognized as a chronic and evolving neurological condition characterized by widespread and heterogeneous structural brain alterations that change over time and contribute to persistent cognitive and functional impairments (Dams-O’Connor et al., 2023; Khattak et al., 2026; Mavroudis et al., 2024; Wilson et al., 2017). However, structural volumetric changes, particularly in relation to recovery process and cognitive functioning continue to be an evolving area of research. In the current study, we used a longitudinal VBM approach to better examine GM volumetric changes across multiple timepoints spanning a 7 – 12-month period, in individuals with msTBI relative to HC, and to evaluate their association with cognitive and functional outcomes. We used a combined approach of longitudinal and cross-sectional analyses, which allowed for a more comprehensive characterization of the brain changes observed following msTBI. Specifically, cross-sectional analyses revealed a spatial distribution of structural alterations, however, they were limited in determining whether such differences remained stable to change over time. To better capture this, longitudinal analyses using a flexible factorial design revealed within-group changes in addition to group and time interactions. Our findings overall demonstrate that changes following TBI reflect a dynamic and diffuse process that also changes over the course of recovery and rehabilitation. Additionally, we observed reductions in subcortical and limbic regions, including the hippocampus, amygdala, thalamus, extending to the pulvinar nuclei, and cerebellar vermis. These regions are involved in memory, emotional regulation, attentional control, and higher-order cognitive integration, suggesting that disruption of distributed subcortical regions may contribute to persistent cognitive and functional difficulties following msTBI.

Contrary to our primary hypothesis, we did not observe significant GM degeneration in the msTBI group over the study period. Global GM, WM, and CSF volumes did not differ between groups at any time point and remained stable across the rehabilitation window. Furthermore, the predicted frontal and temporal cortical reductions were not evident. Instead, most of the subcortical and limbic structures showing significant group-by-time effects were stable or increased in GM volume over time in the msTBI group, with the right amygdala and inferior cerebellar vermis demonstrating a persistent reduction relative to HC. This pattern suggests that within the subacute-to-chronic window captured here, structural change during active rehabilitation may be characterized more by stabilization and partial recovery than by ongoing atrophy. Although, some of this apparent recovery may reflect resolution of injury-related edema and neuroinflammation rather than true tissue gain. In contrast, greater regional GM volume, particularly within the hippocampus and pulvinar, was consistently associated with better cognitive performance, and these brain–behavior associations strengthened at the later timepoint. The absence of progressive atrophy over this interval should be interpreted considering the relatively short follow-up and does not preclude longer-term decline.

One of the most notable findings of the present study was the vulnerability of medial temporal and subcortical structures, particularly the hippocampus and amygdala. Specifically, msTBI exhibited lower right hippocampal GM volume than HC at earlier timepoints, with smaller group differences observed at the third and last timepoint. Although the present study did not demonstrate progressive hippocampal atrophy across all time points, these findings are broadly consistent with prior investigations reporting hippocampal vulnerability following TBI (Ding et al., 2008; Warner et al., 2010). The hippocampus is particularly susceptible to neuroinflammation, hypoxia, and diffuse axonal disruption following trauma, all of which may contribute to impaired memory functioning and long-term cognitive decline. Importantly, greater hippocampal GM volume was consistently associated with stronger verbal memory performance across several CVLT indices in msTBI, supporting the functional relevance of structural preservation within this region during recovery. These findings align with prior studies demonstrating associations between hippocampal integrity and delayed verbal recall following msTBI (Fortier-Lebel et al., 2021; Green et al., 2023; Morrow et al., 2020; Pettemeridou et al., 2025; Till et al., 2008). Similarly, the right amygdala demonstrated persistently lower GM volume in the msTBI group relative to HC across all timepoints. Beyond its established role in emotional processing and affective regulation, the amygdala is highly interconnected with limbic and frontal systems involved in salience detection, stress responsivity, and behavioral regulation (Banks et al., 2007; VanTieghem & Tottenham, 2018). Therefore, structural alterations within this region may contribute to broader cognitive inefficiencies through disruption of limbic-cortical networks. Prior neuroimaging studies have similarly demonstrated degeneration of medial temporal and limbic structures in chronic TBI populations, suggesting these regions may be particularly vulnerable to secondary neurodegenerative processes including neuroinflammation and transneuronal degeneration (Belchev et al., 2022; Bigler, 2013b; Cole et al., 2018; Faden & Loane, 2015; Graham & Sharp, 2019).

The thalamus, extending to the pulvinar nuclei, also emerged as regions of particular interest. The thalamus is consistently implicated in TBI literature due to its role as a central relay hub supporting widespread cortico-subcortical communication networks involved in attention, executive functioning, and memory integration (Cortes et al., 2026; Grossman & Inglese, 2016; Little et al., 2010; Munivenkatappa et al., 2016). In the present study, greater thalamic GM volume was associated with stronger verbal memory performance (CVLT recall) at both baseline and the final timepoint, reinforcing the role of the thalamus in supporting memory-related processing following injury. These findings are consistent with prior studies demonstrating that thalamic injury is associated with widespread cognitive dysfunction and network disconnection after TBI (Little et al., 2010; Scott et al., 2015; Woodrow et al., 2023). Interestingly, msTBI demonstrated consistent longitudinal trajectories in pulvinar nuclei, including relative increases in GM volume across timepoints. Given the pulvinar’s role in attentional allocation and visual-sensory integration (Cortes et al., 2024; Strumpf et al., 2013; Zhou et al., 2016), these findings may reflect adaptive neuroplastic mechanisms or altered reliance on thalamocortical attentional systems involved in salience detection and sensory integration during recovery and rehabilitation (Baird et al., 2024; Munivenkatappa et al., 2016; Sharp et al., 2014). Notably, pulvinar-related cognitive associations were observed primarily in the msTBI group, suggesting that recovery following diffuse injury may involve increased dependence on preserved subcortical attentional networks. Another important finding was the persistent reduction in cerebellar vermis volume in the msTBI group across all longitudinal timepoints. Although commonly associated with motor coordination, the cerebellum is increasingly recognized as contributing to executive functioning, affective regulation, and cognitive processing through extensive cerebello-thalamo-cortical connections (Saalmann et al., 2012; Zhou et al., 2016). Persistent vermian reductions may therefore reflect disruption of broader distributed neural systems involved in cognitive and emotional control. These findings are consistent with prior work demonstrating selective vulnerability of cerebellar and deep subcortical structures following diffuse traumatic injury, particularly in regions susceptible to biomechanical strain and axonal shearing forces (Bigler, 2023).

Importantly, the structural changes observed were meaningfully associated with cognitive performance. Specifically, our findings demonstrated associations between regional brain volumes and cognitive functioning, spanning domains of psychomotor and processing speed, executive functioning, cognitive flexibility, and verbal memory, in individuals with msTBI. Broadly, GM volumes were generally associated with better performance across various measures of cognitive functioning. Notably, subcortical and limbic brain regions, particularly the hippocampus, amygdala, thalamus, and pulvinar nuclei played a prominent role in supporting cognitive performance after TBI. At baseline, greater hippocampal GM volume was associated with better performance across verbal memory, psychomotor and processing speed, cognitive flexibility, and executive functioning, whereas amygdala associations were more limited, confined to long-delay verbal recall. These findings further support the hippocampus’s involvement in memory encoding and retrieval, in addition to the thalamus’s role in facilitating cortico-subcortical communication critical for memory consolidation and executive processes (Mannarelli et al., 2023; Rudolph et al., 2023; Tenório et al., 2025). Among these regions, only the thalamic association with verbal memory was retained and strengthened at the final post-rehabilitation timepoint, whereas hippocampal and amygdala associations were observed primarily at baseline. The baseline associations with cognitive flexibility and executive performance were related to hippocampal rather than thalamic volume, consistent with the thalamus’s broader role as a central hub and sensory relay for distributed cognitive networks (Hwang et al., 2017; Rikhye et al., 2018). Additionally, pulvinar GM volume was associated with better verbal memory performance at baseline. Given the pulvinar’s role in attentional control and visual-sensory integration (Cortes et al., 2024; Strumpf et al., 2013; Zhou et al., 2016), these findings could support a hypothesis for a compensatory mechanism facilitating cognitive performance post-injury and rehabilitation. While the extent of associations between regional volumes and cognitive functioning varied over time, they became more robust at the 6-month timepoint relative to baseline, in correlation with the rehabilitation phase, post-injury.

The current study findings highlight that recovery following msTBI may involve a combination of compensatory neuroplasticity and acute injury-related processes occurring during rehabilitation (Calderone et al., 2024; Hylin et al., 2017; Sharp et al., 2014; Zotey et al., 2023). Although TBI has been associated with neurodegenerative changes (Bigler, 2013a; Faden & Loane, 2015), the present study also demonstrated relative GM volumetric stabilization and improvement in select regions over time, particularly during the rehabilitation phase. These findings suggest that recovery following msTBI may reflect adaptation within distributed neural systems supporting cognitive functioning. Importantly, improvements observed across domains of memory, executive functioning, processing speed, and attention, in conjunction with robust brain-behavior associations in later timepoints, further support the possibility that rehabilitation-related recovery may be associated with measurable brain changes. It is likely that the longitudinal volumetric improvements may reflect partial resolution of injury-related processes such as edema and neuroinflammation (Bigler, 2013a; Faden & Loane, 2015). Thus, these findings highlight the importance of rehabilitation following msTBI, suggesting that targeted interventions may support recovery of distributed cognitive networks (Iaccarino et al., 2015; Rosenbaum et al., 2018; Sharp et al., 2014) (Olsen et al., 2022).

The current study includes several limitations which should be considered when interpreting the present findings. First, the relatively small sample size may limit statistical power and generalizability. Second, variability in injury severity, chronicity, and follow-up intervals may have contributed to heterogeneity in volumetric trajectories across participants. While the longitudinal design improves sensitivity to within-subject change over time, the follow-up remained limited relative to the prolonged neurodegenerative processes that may occur following msTBI. Future investigations using larger cohorts, multimodal imaging approaches, and longer-term longitudinal follow-up will be important for further characterizing the temporal progression of neurodegeneration and its relationship to cognitive and functional outcomes following injury.

## Data Availability

The authors will share the data with qualified investigators whose proposal of data use had been approved by an independent review committee. Further inquiries may be directed to the corresponding author.

## Data availability

The original contributions presented in this study are included in this article/supplementary material, further inquiries may be directed to the corresponding authors.

## Ethics statement

Ethical approval and consent to participate: All data presented in this study were collected upon approval from Advarra’s Institutional Review Board.

## Conflict of Interest

The authors declare that the research was conducted in the absence of any commercial or financial relationships that may be construed as a potential conflict of interest.

## Funding

This work was supported by the Synaptics Foundation, Bakersfield CA

## Author Contributions

RJ, BBO, JY, JA, and GG made substantial contributions to conceptualization and drafting of the manuscript. RJ and BBO contributed towards statistical analyses. RJ, BBO, JA, and GG contributed towards data collection and data analyses. All authors participated in manuscript revisions and have given final approval of this version.

## Acknowledgments

Special thanks to the participating families and staff who contributed toward data collection.

## Notes

### Competing Interest Statement

The authors have declared no competing interest.

### Author Declarations

All participants provided written informed consent and Advarra Institutional Review Board gave ethical approval for this study.

### Summary of Updates

Table 3 updated, Table 1 updated to clarify study timeline

